# Development and Validation of a parsimonious AI-Based Risk Score for Mortality in Heart Failure: A UK cohort study

**DOI:** 10.1101/2025.09.29.25335268

**Authors:** Nouman Ahmed, Shishir Rao, Nathalie Conrad, Malgorzata Wamil, Ben Omega Petrazzini, Zhengxian Fan, Guyu Zeng, Jie Lian, Kazem Rahimi

## Abstract

**Background:** Accurate risk stratification in heart failure (HF) is crucial to guide clinical decisions, optimise therapeutic strategies and inform resource allocation. Existing widely used tools like from the Meta-Analysis Global Group in Chronic Heart Failure (MAGGIC) show modest discriminatory performance and rely on specialised tests (e.g., echocardiography), while advanced Artificial Intelligence (AI) models, though more accurate, face significant barriers to adoption and integration into routine care pathways due to their complexity and data access issues. By leveraging insights from a complex AI model and feature engineering, we aim to develop and validate a both, parsimonious and high-performing, AI-based risk model for all-cause mortality in HF patients, specifically designed for easy interpretation and accessibility in a clinical setting.

**Methods:** Using a cohort of 373,389 patients with HF (≥18 years; 1,153 English practices for derivation, 289 for validation) from the Clinical Practice Research Datalink (CPRD) Aurum dataset, we developed and validated an AI-based risk prediction model for all-cause mortality. An initial Multi-layer Perceptron (MLP) model with a survival framework was trained with variables from the MAGGIC score. This MLP model was then enhanced by incorporating highly predictive by incorporating highly predictive comorbidity features identified through an explainable Transformer model trained on longitudinal electronic health record (EHR) data and subsequently distilled through a novel feature engineering approach to derive a final, optimised 11-variable model (named ‘SIMPLE-HF’ - **S**implified **I**ntelligent **M**ortality **P**rediction for **L**ongitudinal **E**HRs in **HF** patients). The resulting model utilised readily available variables at the point-of-care, including age, BMI, year of birth (as a proxy for birth cohort effects), and key comorbidities such as cancers. We examined the discrimination and calibration of SIMPLE-HF on the validation dataset and compared performance against a MAGGIC score adapted for use on routine EHR (termed as MAGGIC-EHR). In secondary analyses, we investigated the performance of SIMPLE-HF for the prediction of major adverse cardiovascular events and hospitalisation.

**Findings:** The SIMPLE-HF model demonstrated significantly improved discriminatory performance (C-index: 0.801, 95% CI [0.795, 0.806]) compared to the benchmark MAGGIC-EHR Cox model (0.735, [0.728, 0.741]), while maintaining acceptable calibration. Clinical impact analysis further revealed that SIMPLE-HF consistently captures more true events while flagging fewer patients as high-risk. For every 1000 patients screened, at a high-risk threshold of 0.60, SIMPLE-HF identifies 199 patients who will have an event, whereas MAGGIC-EHR captures only 99 true events. Similarly, SIMPLE-HF achieved improved performance on cardiovascular events and hospitalisation prognostication as compared to the benchmark model.

**Interpretation:** Leveraging insights from a complex EHR-trained AI model into easily collected clinical features, we enhance the accuracy and practicality of HF mortality risk prediction. This novel approach offers a pathway to clinically implementable tools that balance predictive accuracy with pragmatic utility and has the potential to improve HF risk stratification, along with offering significant cost efficiency by removing the need of specialised tests.

**Research in context:** *Evidence before this study:* Before this study, the risk stratification of patients with heart failure (HF) presented a clear challenge. Existing clinical risk scores, such as the widely known MAGGIC score, demonstrate only modest predictive accuracy (typically with C-indices between 0.61 and 0.75) and often depend on variables from specialised tests like echocardiography, which are not always available in routine care, particularly in primary care settings. This limits their practical utility. In contrast, advanced artificial intelligence (AI) models, such as those using Transformer architectures trained on extensive longitudinal electronic health records, have shown superior predictive performance (C-indices > 0.80). However, their complexity, reliance on vast datasets, and significant computational requirements create major barriers to their adoption and integration into everyday clinical workflows. On Feb 1, 2025, we searched PubMed for English-language studies published between Jan 1, 2015, and Jan 1, 2025, using the terms “heart failure”, “risk prediction”, “mortality”, “prognostic model”, and “machine learning”. Our search did not identify any parsimonious risk models that achieved high discrimination and were specifically validated on a large, representative UK cohort using only readily available clinical data.

*Added value of this study:* This study introduces and validates SIMPLE-HF, a parsimonious, 11-variable risk score for all-cause mortality in HF patients that directly addresses the performance-practicality gap. By using a novel “distillation” methodology, we translated the predictive insights from a complex, high-performing Transformer AI model into a simple, interpretable model. Our model achieves significantly higher discrimination (C-index: 0.801) for all-cause mortality compared to an EHR-adapted benchmark model (MAGGIC-EHR C-index: 0.735). Crucially, SIMPLE-HF relies exclusively on variables that are routinely collected at the point-of-care, eliminating the need for specialised tests. Furthermore, the model demonstrates superior clinical impact; for every 1000 patients screened at a 60% risk threshold, SIMPLE-HF correctly identifies 199 patients who will have an event, compared to only 99 identified by the benchmark model

*Implications of all the available evidence:* The combined evidence suggests that SIMPLE-HF is a robust and practical tool that can be immediately implemented in diverse clinical settings, including primary care, to improve the accuracy of heart failure risk stratification. For clinical practice, it offers a more accessible and precise method to guide therapeutic decisions and resource allocation without requiring additional specialised tests. For future research, our distillation methodology provides a new paradigm for developing clinically implementable prediction tools. This approach harnesses the power of complex AI to create simple, transparent, and effective models, a strategy that could be applied to improve risk prediction across numerous other medical conditions. External validation of the SIMPLE-HF model in diverse international cohorts is a recommended next step to confirm its generalisability.

## Introduction

Heart failure (HF) represents a significant global public health challenge, with high rates of hospitalisation and mortality.^1^ Effective risk stratification is paramount in HF management to guide therapeutic decisions, allocate healthcare resources efficiently, and inform patient prognosis, especially as the projected population with HF in the UK is expected to double by 2040.^2^ While numerous prognostic tools exist, including the Meta-Analysis Global Group in Chronic Heart Failure (MAGGIC) score^3^, they often present modest discriminatory power and necessitate specialised investigations (such as echocardiography for estimation of ejection fraction). Such limitations likely contribute to their underutilisation in routine clinical practice, despite guideline recommendations underscoring the need for accurate, yet practical and parsimonious, prognostic models.^4,5^

Over the past decade, access to extensive healthcare datasets, including electronic health records (EHRs), has expanded considerably.^6,7^ These rich data sources offer comprehensive, longitudinal capture of patient health trajectories, providing unprecedented opportunities to unravel the complex interplay of factors influencing chronic diseases like HF^8-11^ Concurrently, sophisticated artificial intelligence (AI) methodologies, notably Transformer-based architectures, have demonstrated superior discrimination and calibration in predicting clinical outcomes by modelling complex, latent associations within extensive longitudinal EHR data.^12,13^

However, the advanced capabilities of these state-of-the-art AI models present their own set of challenges. Their reliance on rich, longitudinal datasets and substantial computational resources often restricts their applicability in settings with limited data availability or infrastructure.^14-16^ This gap highlights a critical unmet need: to harness the predictive power demonstrated by complex AI systems without inheriting their barriers to implementation.

To address these limitations, we aimed to balance predictive accuracy with ease of use, thereby offering a novel pathway to improve risk stratification in diverse clinical settings. We derived and validated an AI-based parsimonious risk score model. The generalisability of this approach was also explored through validation on secondary outcomes, including rehospitalisation and subsequent major cardiovascular events.

## Methods

### Overview

#### Data Source and validation strategy

We used anonymised EHR data from the Clinical Practice Research Datalink (CPRD) Aurum and its linked datasets. CPRD provides detailed patients’ records from primary care, including demographics, diagnoses, prescribed HF therapy, and health-related lifestyle variables, on approximately 20% of the UK population and is broadly representative of age, sex, ethnicity, geographical spread, and socioeconomic deprivation.^17^ Linked data is available for a subset of English practices, covering about 50% of all CPRD records, and further provides information on hospital admissions and death certificates, from Hospital Episode Statistics Admitted Patient Care (HES-APC) and Office for National Statistics (ONS) respectively.^17^

We included data from 1,442 contributing GP practices in England. To ensure robust validation, a subset of practices was randomly assigned to the derivation cohort for model development (80% - 1,153 practices), and the remaining to the validation cohort (20% - 289 practices). This practice-level split mitigates data leakage by ensuring the model is tested on patients from practices it has never seen before. This method provides a more rigorous assessment of model generalisability than splitting by individual patients, which can artificially inflate performance by testing the model on the same practice-specific data patterns it was trained on (details in **Supplementary Methods: Clarification on CPRD validation study**).

#### Cohort Selection

Included in the study were men and women registered with their general practice for at least one year during the study period (01/01/2010 to 31/12/2020) whose records were classified by the CPRD as acceptable for use in research and approved for HES and ONS linkage. Among these, we identified an open cohort of individuals aged 18 years or older with incident HF. We identified HF diagnoses using established phenotyping algorithms for CPRD Aurum,^10^ considered diagnoses in primary care or secondary care records and regarded each patient first record of HF as their incident diagnosis. We excluded individuals who had a HF diagnosis before the start of the study (01/01/2010), before their 18^th^ birthday, or within the first 12 months of registration with their general practice.

Each patient was followed up starting at the latest of the following: January 1st 2010, their 18th birthday, 12 months after GP registration. Follow-up ended at the earliest of the following: date of outcome of interest (i.e., mortality), last collection date from practice, patient’s last date in practice, or December 31st, 2020. For each HF patient included in the study, the index date (i.e., baseline), was randomly selected from the period between first HF diagnosis and end of study-period. This approach simultaneously ensures capture across the spectrum of calendar years and ages, and better reflects the reality of clinical practice, where patients are assessed at various stages of their care journey.^18^

#### Primary Outcome Definition

The primary outcome was all-cause mortality, at 12 and 36 months, determined from linked ONS mortality records.

#### Analyses of secondary outcomes

To explore the broader utility of the our approach, supplementary analyses were conducted to assess the performance in predicting two additional clinically relevant outcomes. These included (1) major adverse cardiovascular event, defined as a composite of fatal or non-fatal ischaemic heart disease, myocardial infarction, transient ischaemic attack, or stroke, and (2) any hospitalisation after incident HF. These outcomes were identified using both previously published and our own curated code dictionaries.^10,19^

#### AI Development Framework

The AI model development was an iterative process, with each step building upon the last **(Figure S2)**. All models were developed using a multi-layer perceptron (MLP) within a discrete-time survival framework using Ordinary Differential Equations Networks (SODEN).^20^

We began by developing a model using the established MAGGIC predictor set to serve as a benchmark for subsequent enhancements. To improve predictive power, this feature set was then augmented based on findings from a more complex, explainable Transformer model, which highlighted the prognostic value of previously under-appreciated variables like specific cancer histories and renal failure (**Table S2** for the exact explainability variables and their prevalence in the CPRD cohort).^12^ We also incorporated the patient’s year of birth as a proxy for birth cohort effects (i.e., generational differences in lifestyle, healthcare, and environmental exposures that can influence health outcomes), ^21-23^ creating an enriched intermediate model with 21 variables. As a final step to enforce parsimony, we used SHapley Additive exPlanations (SHAP) to analyse and distil these 21 features down to only the most impactful predictors. This process resulted in our final model, SIMPLE-HF (**S**implified **I**ntelligent **M**ortality **P**rediction for **L**ongitudinal **E**HRs in **HF** patients), which uses a refined set of just 11 variables: year of birth, occurrence of HF 18 months before baseline, age, cancer history, prescription of beta-blockers or ACE inhibitors/ARBs, body mass index (BMI), history of acute renal failure, pneumonia, cardiac arrest, respiratory failure, and chronic obstructive pulmonary disorder (**Figure S3** for results of SHAP analysis).

Details on how the individual features were extracted and details regarding modelling can be found in **Supplementary Methods: Feature Selection, AI Modelling, and SHAP Analyses, Figure S2-S3**.

#### Benchmark modelling

For comparative purposes, the MAGGIC score was adapted for application to EHR data (termed MAGGIC-EHR), based on previously published methodology.^3^ All predictors from the original MAGGIC score were extracted from EHR data, except for Left Ventricular Ejection Fraction (LVEF), which is infrequently recorded in large-scale primary care EHR data. As a practical substitute for LVEF, we incorporated a categorical variable representing HF subtype (i.e., reduced, preserved, or unknown LVEF), derived using established phenotyping codes.^25^ To address data missingness, imputation was separately carried out on derivation and validation datasets^26^ (details in **Supplementary Methods: Implementation of MAGGIC-EHR models)**.

#### Performance Analyses

We evaluated models’ performance on the UK validation dataset using multiple metrics. For discrimination, we assessed the concordance index (C-index) and area under the precision-recall curve (AUPRC). Model calibration was assessed graphically, by comparing the agreement between the observed and predicted risk using a smoothed calibration curve fit using restricted cubic splines with three knots, and quantitatively by calculating the integrated calibration index (ICI) metric.^27^ In addition to this, we also conducted net benefit analyses at clinically relevant thresholds.

For all analyses, individual patient risk scores were derived by estimating survival probabilities over a 60-month follow-up period, focusing on the 12-, and 36-month timepoint. This is a characteristic of the SODEN model, which is able to estimate the survival probability at every month in the follow-up period.^20^

Additionally, we conducted discrimination analyses in subgroups defined by sex, age, baseline systolic blood pressure, comorbidities and medication use at baseline.

Calibration and net benefit analyses for 12 and 36-month prediction were also conducted for all models to assess model performance at different follow-up durations.

Furthermore, we conducted an impact analysis to evaluate true positive and negative capture at the 50% decision threshold of the SIMPLE-HF and MAGGIC-EHR models on UK validation data in line with research recommendations by HF clinical guidelines.^5^

## Results

### Population Characteristics

Among 19,094,480 individuals contributing to data and aged over 18 years between 01/01/2010 and 31/12/2020, we identified 373,389 patients with incident HF (**Figure S1**) for analysis (77,712 patients in validation) with median follow-up of 7 months (interquartile interval [IQI]: [2, 20]). The median age was 80 years (IQI: [70,87]) and approximately, one-third of the cohort had a history of diabetes, stroke, or myocardial infarction along with more than half of the cohort being prescribed beta-blockers.Approximately, 42% of patients died in five years following baseline.

### Analyses

SIMPLE-HF achieved higher C-index (0.801; 95% CI [0.795,0.806]) than MAGGIC-EHR (0.735; [0.728,0.741]) along with higher AUPRC (0.684 [0.677,0.691] vs 0.560 [0.553, 0.567]) (**Figure 1, S4, Table S3**). SIMPLE-HF also showed less deviation from overall performance within sub-groups (**Figure 1**). Both models were well calibrated as shown with their closeness of their calibration curves to the reference curve and low ICI estimates (**Figure 2A, Tables S4**) Decision curve analysis showed that SIMPLE-HF provided significantly greater net benefit than other strategies across the spectrum of clinically relevant thresholds (up to ∼0.6) (**Figure 2B**). Analyses investigating 36-month predictions showed similar discrimination and calibration estimates (**Figure S5, Table S5**).

**Figure 1.**
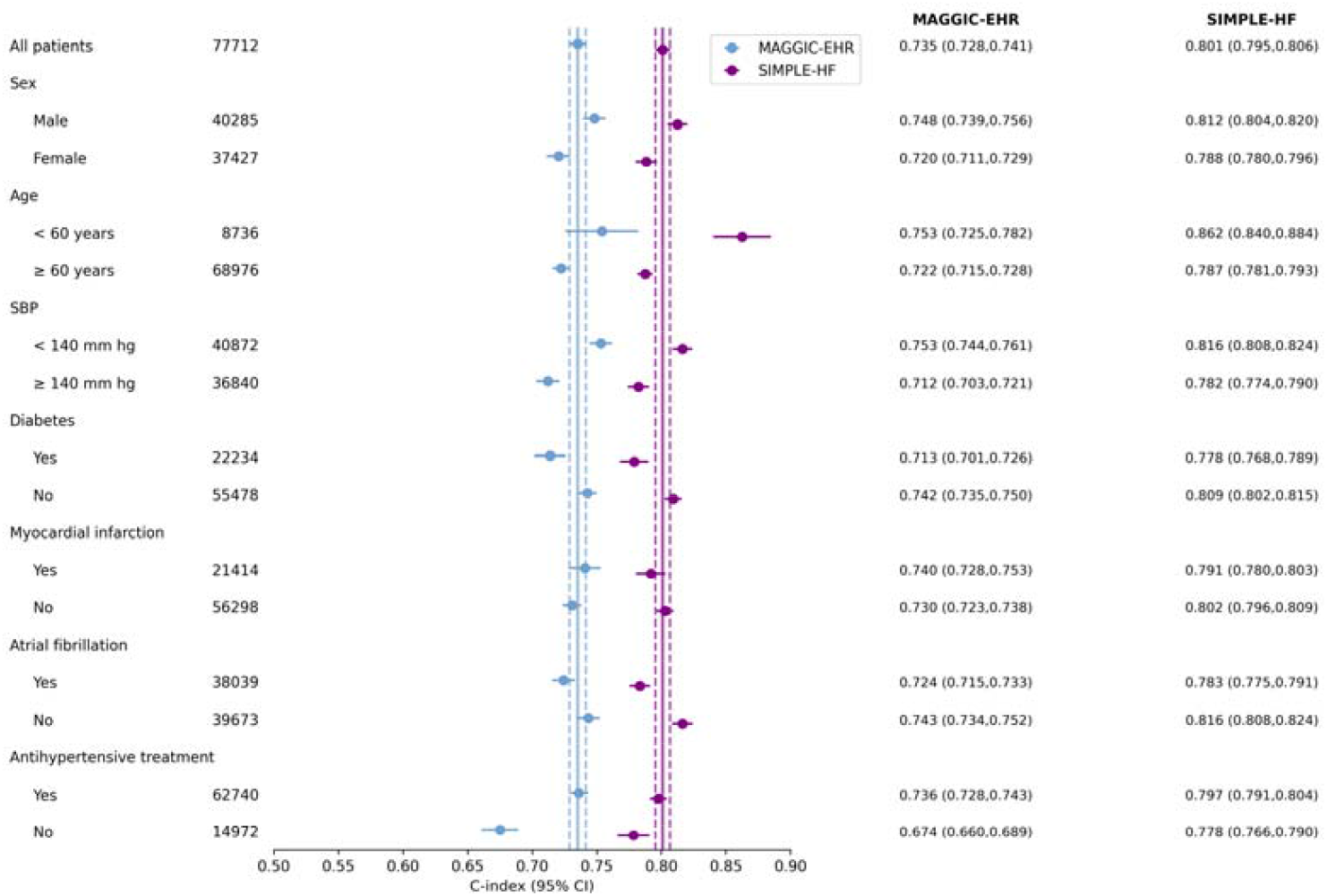
Models’ discrimination by concordance index (C-index) and associated 95% confidence intervals (CI) in overall cohort and subgroups for 12-month all-cause mortality risk prediction on UK validation data. Blue and Purple lines represent C-index on “all patients” in the validation cohort for MAGGIC-EHR and SIMPLE-HF respectively. These lines are provided to visually demonstrate deviation in subgroup discrimination performance from overall cohort performance for each model

**Figure 2:**
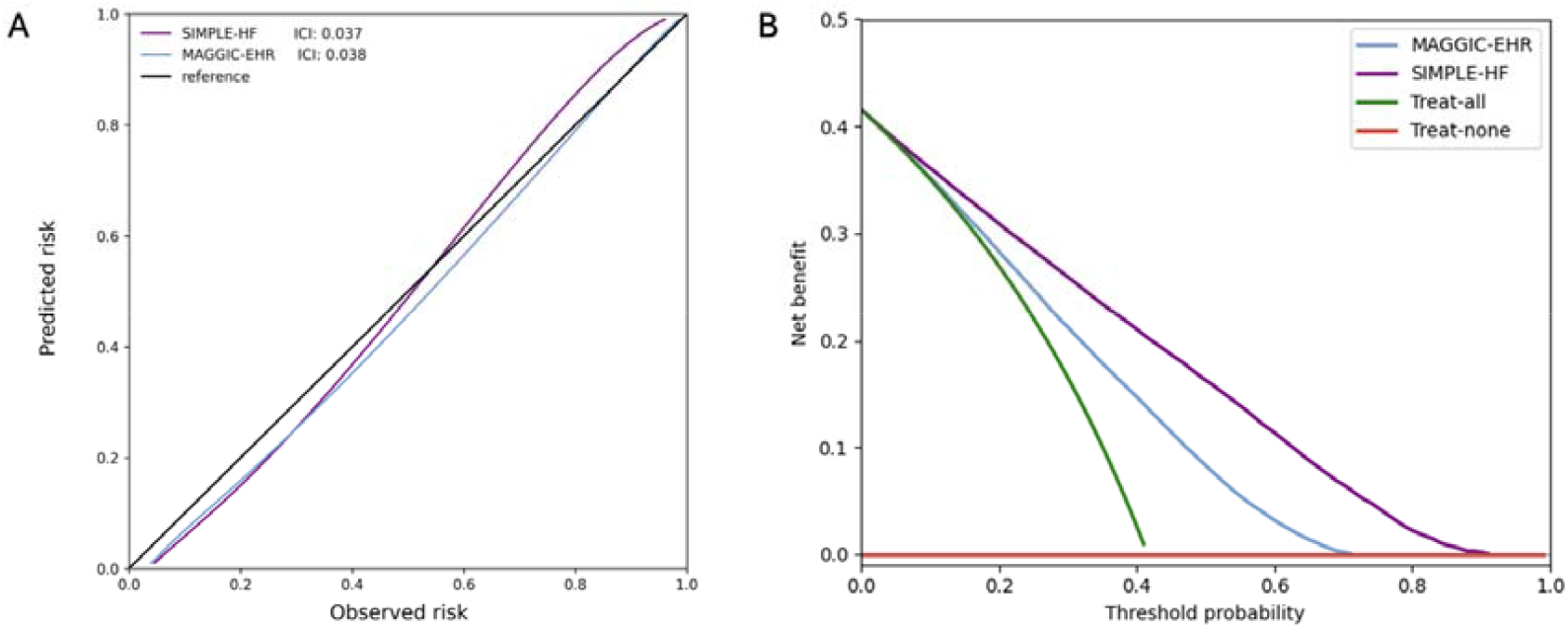
Calibration curves, and net benefit analysis for 12-month risk prediction of all-cause mortality in UK validation data. Calibration curves with integrated calibration indices (ICI) (A), and decision curve analysis (B) are presented for all-cause mortality at 12-month timepoint. For (A) ICI, lower is better with reference (black line) presenting optimal ICI of 0.0.

**Table 1.**
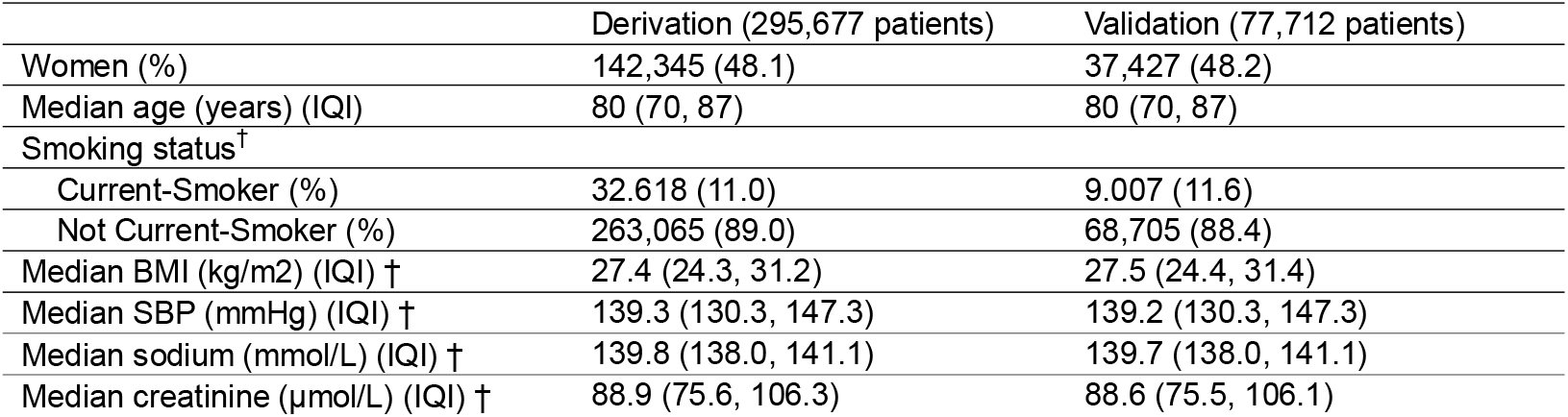
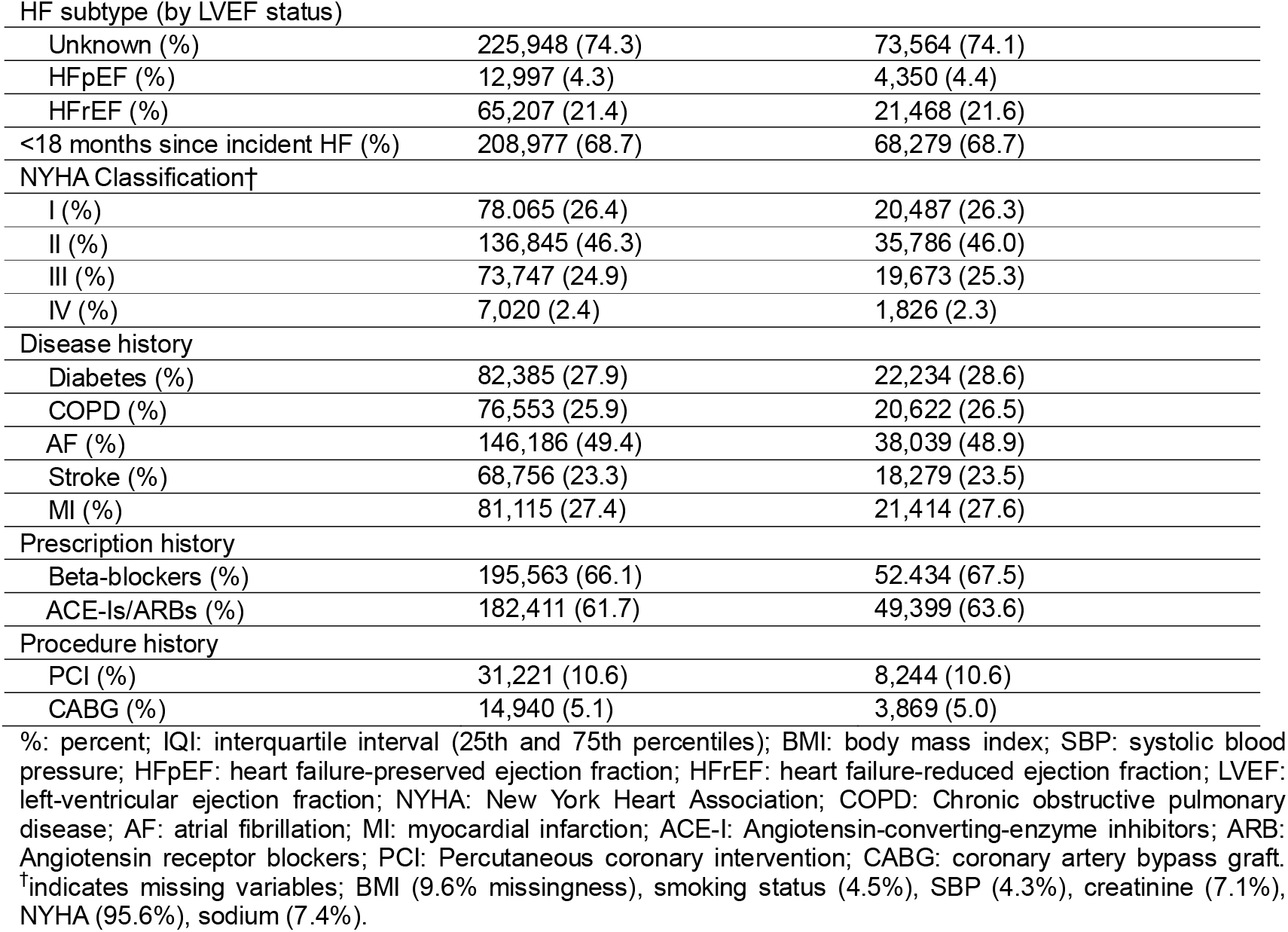
Population characteristics for derivation and validation datasets in UK data study at baseline.

|  | Derivation (295,677 patients) | Validation (77,712 patients) |
| --- | --- | --- |
| Women (%) | 142,345 (48.1) | 37,427 (48.2) |
| Median age (years) (IQI) | 80 (70, 87) | 80 (70, 87) |
| Smoking status<sup>†</sup> |  |  |
| Current-Smoker (%) | 32.618 (11.0) | 9.007 (11.6) |
| Not Current-Smoker (%) | 263,065 (89.0) | 68,705 (88.4) |
| Median BMI (kg/m<sup>2</sup>) (IQI) † | 27.4 (24.3, 31.2) | 27.5 (24.4, 31.4) |
| Median SBP (mmHg) (IQI) † | 139.3 (130.3, 147.3) | 139.2 (130.3, 147.3) |
| Median sodium (mmol/L) (IQI) † | 139.8 (138.0, 141.1) | 139.7 (138.0, 141.1) |
| Median creatinine (μmol/L) (IQI) † | 88.9 (75.6, 106.3) | 88.6 (75.5, 106.1) |
| HF subtype (by LVEF status) |  |  |
| Unknown (%) | 225,948 (74.3) | 73,564 (74.1) |
| HFpEF (%) | 12,997 (4.3) | 4,350 (4.4) |
| HFrEF (%) | 65,207 (21.4) | 21,468 (21.6) |
| <18 months since incident HF (%) | 208,977 (68.7) | 68,279 (68.7) |
| NYHA Classification† |  |  |
| I (%) | 78,065 (26.4) | 20,487 (26.3) |
| II (%) | 136,845 (46.3) | 35,786 (46.0) |
| III (%) | 73,747 (24.9) | 19,673 (25.3) |
| IV (%) | 7,020 (2.4) | 1,826 (2.3) |
| Disease history |  |  |
| Diabetes (%) | 82,385 (27.9) | 22,234 (28.6) |
| COPD (%) | 76,553 (25.9) | 20,622 (26.5) |
| AF (%) | 146,186 (49.4) | 38,039 (48.9) |
| Stroke (%) | 68,756 (23.3) | 18,279 (23.5) |
| MI (%) | 81,115 (27.4) | 21,414 (27.6) |
| Prescription history |  |  |
| Beta-blockers (%) | 195,563 (66.1) | 52,434 (67.5) |
| ACE-Is/ARBs (%) | 182,411 (61.7) | 49,399 (63.6) |
| Procedure history |  |  |
| PCI (%) | 31,221 (10.6) | 8,244 (10.6) |
| CABG (%) | 14,940 (5.1) | 3,869 (5.0) |

%: percent; IQR: interquartile interval (25th and 75th percentiles); BMI: body mass index; SBP: systolic blood pressure; HFpEF: heart failure-preserved ejection fraction; HFrEF: heart failure-reduced ejection fraction; LVEF: left-ventricular ejection fraction; NYHA: New York Heart Association; COPD: Chronic obstructive pulmonary disease; AF: atrial fibrillation; MI: myocardial infarction; ACE-I: Angiotensin-converting-enzyme inhibitors; ARB: Angiotensin receptor blockers; PCI: Percutaneous coronary intervention; CABG: coronary artery bypass graft. †indicates missing variables; BMI (9.6% missingness), smoking status (4.5%), SBP (4.3%), creatinine (7.1%), NYHA (95.6%), sodium (7.4%).

In impact analysis, SIMPLE-HF increases true positives by 35% (4,218 patients) while reducing false negatives by 36% (4,818 patients) for 12-month prediction as compared to MAGGIC-EHR (**Figure 3; Table S6**). Higher true positives and fewer false negatives directly translated into SIMPLE-HF outperforming MAGGIC-EHR in both PPV and sensitivity, showing increases of 0.072 and 0.067 for 12-month prediction. Similar results were found for 36-month prediction **(Figure S6; Table S6)**.

**Figure 3.**
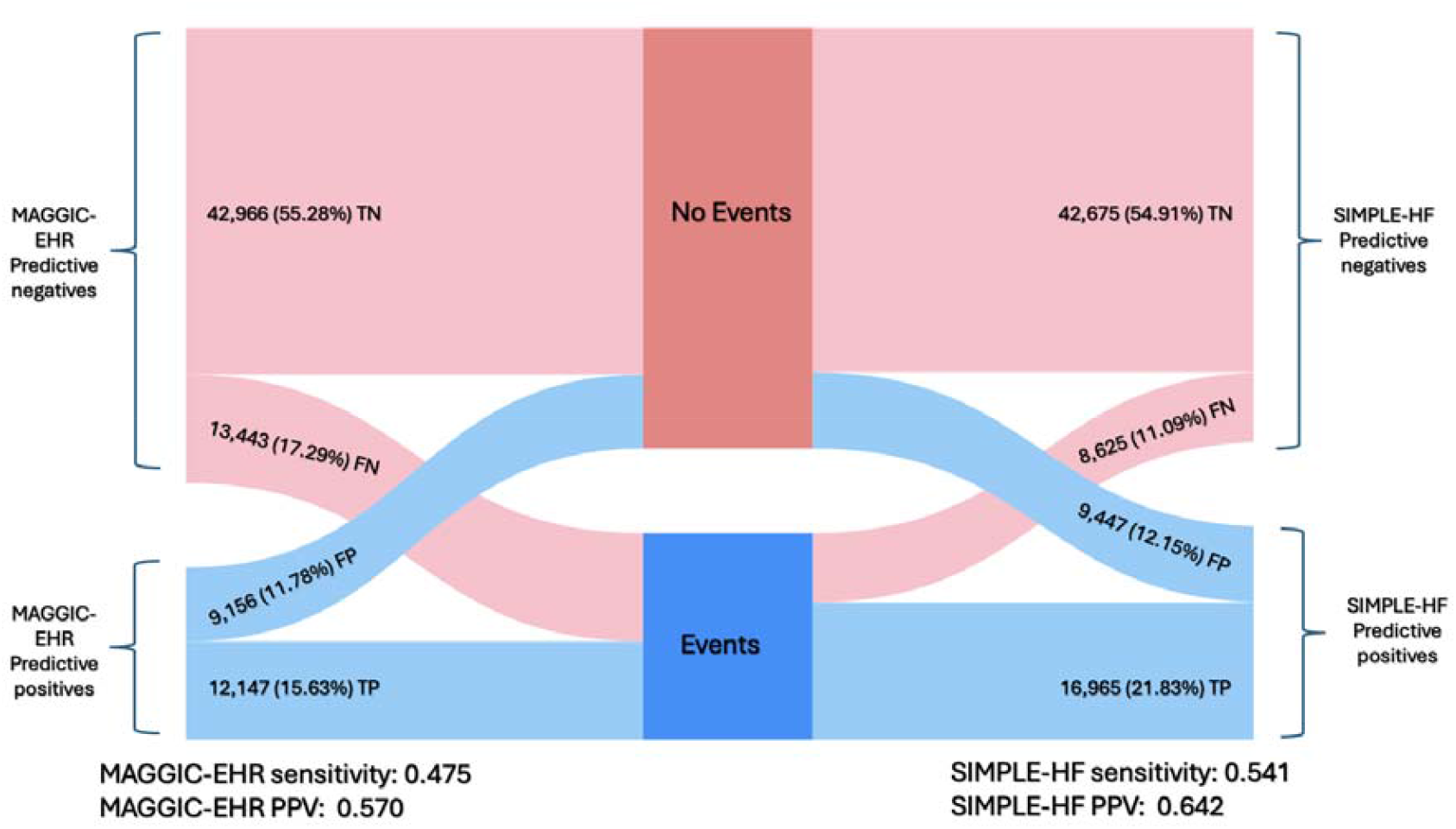
Impact analyses at the 50% decision threshold for 12-month all-cause mortality prediction on UK validation data. Sankey diagrams compare predicted outcomes between the models, showing how patient classification compares to actual outcomes, “Events” (i.e., death) and “No events” categories at 50% threshold (denoted as dark blue and red respectively). TP: true positive; TN: true negative; FP: false positive; FN: false negatives.

The clinical-impact curves (**Figure 4**) illustrate the number of patients, per 1000 screened, who would be classified as high-risk and who would subsequently experience the outcome across a range of decision thresholds. SIMPLE-HF consistently outperforms MAGGIC-EHR: it marks fewer patients as high-risk yet still captures more of those who go on to have the event. The contrast is most striking at higher clinically relevant thresholds, such as 0.60, where SIMPLE-HF identifies 199 true events among 257 flagged patients, while MAGGIC-EHR captures only 99 events among 145 flagged patients—half the true event capture of SIMPLE-HF. This pattern holds at every threshold examined, demonstrating superior clinical yield with fewer unnecessary alerts. The results persisted for 36-month prediction as well (**Figure S7**).

**Figure 4.**
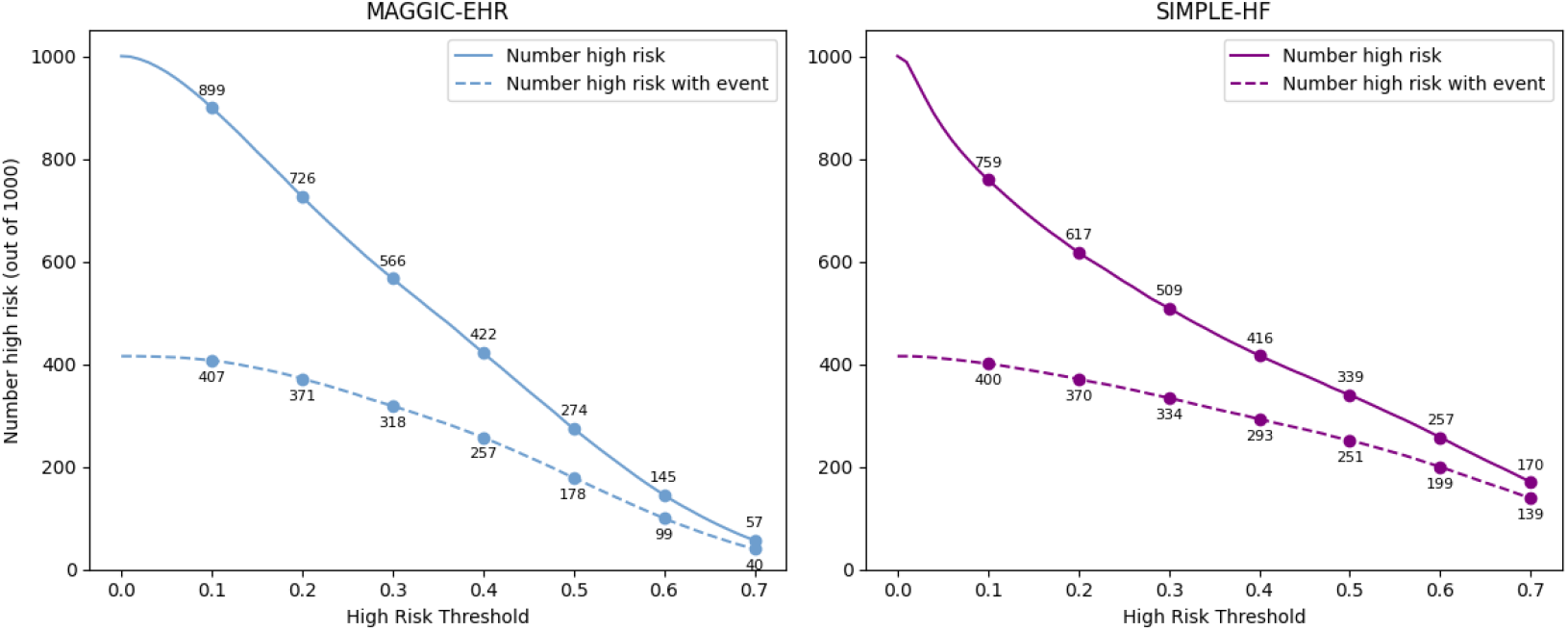
Clinical-impact curves for 12-month all-cause mortality prediction in the UK validation cohort. Comparison of MAGGIC-EHR (left) with the SIMPLE-HF model (right). For each possible decision threshold on the x-axis, the solid line represents the number of patients per 1000 who would be classified as high risk, whereas the dashed line represents the subset of those high-risk patients who experienced the outcome (death). Numeric labels denote the corresponding absolute counts

### Analyses of secondary outcomes

In addition to our primary outcome of all-cause mortality, we conducted analyses on secondary outcomes for the prediction fatal and non-fatal cardiovascular outcomes (median follow-up of 4 months - IQI: [1, 14]) and hospitalisation (median follow-up of 4 months IQI: [1, 12]). These secondary outcomes analyses showed similar estimates of discrimination and calibration and SIMPLE-HF out-performed MAGGIC-EHR, for 12-, and 36-month prediction (**Figure S8-S12, Tables S3, S5, S6**).

## Discussion

In this study, we successfully developed and validated a parsimonious, 11-variable risk score that achieves high discrimination (C-index: 0.801, 95% CI [0.796–0.806]) for predicting all-cause mortality in patients with HF. Importantly, the model, relies exclusively on variables readily available at the point-of-care, hence removing the need for specialised tests or longitudinal data streams.

The existing landscape of HF prognostication tools presents a dichotomy. Conventional risk scores, such as MAGGIC or the Seattle Heart Failure Model typically yield C-indices in the range of 0.61 to 0.75 ^3,28^ These models have limited clinical utility in routine care settings due to their modest discriminatory properties and reliance on results of specialised testing procedures. This limitation is particularly significant since much of HF care in the UK and other countries occurs in primary care.^29,30^ Additionally, utilising these risk scores on HF patients throughout their care journey requires model updates with new variables, necessitating repeated specialised testing (as in requiring subsequent echocardiography for LVEF measurements). Conversely, recent cutting-edge AI models leveraging dense, longitudinal EHR data can achieve C-indices exceeding 0.8.^12^ However, their substantial data and computational demands restrict their widespread adoption and integration at the point-of-care.^14-16^

The present work directly addresses this performance-practicality gap. Using a novel approach, we ‘distilled’ insights from a sophisticated Transformer-based AI model, initially trained on rich longitudinal EHR data, into a simpler MLP-based model. This process, guided by SHAP-based feature selection, began with verifying that an MLP architecture using standard MAGGIC predictors could outperform the traditional Cox-based MAGGIC model. The subsequent augmentation with Transformer-identified comorbidity signals further surpassed with **(Figure S4)**. Lastly, feature distillation yielded the final 11-variable SIMPLE-HF model (∼ΔC-index +0.07 as compared to MAGGIC-EHR). This model not only significantly surpassed the performance of an EHR-adapted MAGGIC benchmark but also substantially reduced the data collection burden by eliminating the need for variables like LVEF, which require specialised investigations. Moreover, the SIMPLE-HF includes variables related to co-morbidities which is in-line with the current research that complex multimorbidity, rather than HF itself, contributes to over 40% of mortality in HF patients.^8,31,32^ This highlighting a significant gap in current risk assessment approaches, which SIMPLE-HF fulfils.

Beyond its immediate clinical relevance for the management of HF patients, this study also carries important methodological implications for the development of clinical prediction models. It showcases a pragmatic approach to harness the signal-finding capabilities of complex AI architectures, to inform the development of simpler and more implementable tools suitable for routine clinical use. Our findings suggest that explainable AI techniques can serve not only as ‘black box’ models for complex modelling paradigms, but also as powerful knowledge discovery and translation tools. Ultimately, this approach has the potential to bridge the gap between the exploratory power of advanced AI and the pragmatic requirements of clinical decision support at the bedside.

The clinical utility of the SIMPLE-HF score is manifold. Firstly, all 11 input variables are routinely collected during clinical encounters and require no specialised imaging, assays, or continuous longitudinal monitoring. In fact, they can also be compiled in the form of a checklist. Secondly, when using higher risk thresholds that are more relevant to clinical practice, the SIMPLE-HF model identifies more than twice as many patients who go on to have events, compared to existing models. Third, our model is validated for prognostication across the entire HF trajectory, rather than being limited to the time of initial diagnosis. By employing random baseline selection, the model avoids bias toward any specific disease stage - whether early or advanced. This makes it more applicable to real-world clinical practice, where patients often present at varying points along the disease continuum. Collectively, these attributes suggest that the SIMPLE-HF score could be deployed even in healthcare settings with limited EHR interoperability, thereby supporting broader and more equitable access to robust risk stratification tools.

This study has several notable strengths. These include the use of a large, nationally representative primary care dataset linked to hospitalisation and mortality records, a rigorous validation strategy employing a practice-level split, and the novel application of a distillation approach to develop a clinically practical tool. The significant improvement in discrimination over established benchmarks with reduced data requirements underscores the potential of our methodology.

However, certain limitations warrant consideration. Firstly, while CPRD Aurum is representative of the English population, further external validation in diverse international cohorts and healthcare systems is necessary to confirm generalisability of our approach in other settings. Secondly, our adaptation of the MAGGIC score for EHR data (MAGGIC-EHR), while necessary and pragmatic to allow its implementation using routine clinical data, represents a modification of the original score with several variables (e.g. NYHA or LVEF) missing to a significant extent in electronic health records, which should be considered when interpreting comparisons. Lastly, our model’s features were optimised for predicting all-cause mortality and its application to secondary outcomes should therefore be viewed as exploratory for now. To create optimised models for secondary outcomes, our methodology could be repeated by training the complex AI specifically for each of those endpoints and then distilling a new set of key predictors.

## Conclusion

By translating the predictive power of a complex Transformer model into a streamlined, 11-variable risk score, this study offers a robust, clinically actionable tool for stratifying mortality risk in HF patients, bridging the gap between advanced AI and real-world clinical implementation. This approach not only provides an immediately applicable risk stratification instrument but also exemplifies a broader paradigm for translating the sophisticated analytical power of advanced AI into tangible, accessible, and impactful clinical decision support systems, ultimately aiming to improve patient outcomes.

### Contributors

NA and SR conceptualised and designed the study. KR and SR provided supervision for the project. NA conducted all quantitative analyses. NA and SR wrote the first draft of the manuscript. NA, SR, NC, BOP, GW, ZF, GY, KR, and JL provided methodological insights on the manuscript. All authors were involved in revising the manuscript. NA and SR were responsible for the decision to submit the manuscript for publication.

## Data sharing

This study was approved by the CPRD Independent Scientific Advisory Committee (protocol number 20_095). The CPRD data used in this study are available to researchers through a licence agreement following protocol approval from the Independent Scientific Advisory Committee. These data are not publicly available due to licensing restrictions. Details about data access and sharing policies can be found at the CPRD website.

## Data Availability

Clinical Practice Research Datalink (CPRD) Aurum dataset was used which is available by application.

