## Supplementary Methods, Tables, and Figures for "Development and Validation of a parsimonious AI-Based Risk Score for Mortality in Heart Failure: A UK cohort study"

#### Clarification on CPRD validation study

CPRD data was divided into derivation and validation datasets by selecting patients from different general practices (GPs). Derivation dataset was used for model training while the validation data served as validation for the trained models. For MLP models, 5% of the derivation dataset was randomly selected to conduct end-of-epoch testing. This was done to avoid overfitting, which is a concern in AI parametric modelling. This testing, done at the end of every epoch (i.e., one full iteration of training in which the model has trained on all patients selected for training), ensured that the model is not overfitting on the derivation dataset (i.e., ultimately aiming to avoid data “memorisation”). After model training was estimated to have converged on the derivation dataset, the AI models were evaluated on the validation dataset. All patients in the derivation dataset were used for the fitting of statistical cox models (i.e., MAGGIC-EHR models).

#### Feature Selection

The initial features included the ones in the MAGGIC model. These included sex, age, smoking status, diabetes, systolic blood pressure, creatinine, body mass index (BMI), New York Heart Association (NYHA) Functional Class, chronic obstructive pulmonary disease, any use of beta-blockers, any use of ACE inhibitor/ Angiotensin receptor blockers, incident HF diagnosis ≥18 months prior to baseline, and HF subtype (i.e., preserved, reduced, or unknown according to previously published method^1^).

To enhance this feature set, we included knowledge learned from more complex model and data, namely a Transformer-based survival framework trained on large-scale longitudinal EHR data.^2^ The authors identified top ten encounters (diagnosis, medications, and procedures) that were the most predictive of a higher risk by the transformer model. This essentially included history of co-morbidities, namely, cardiac arrest, secondary malignant neoplasm of lung, hepatic failure, respiratory failure, pneumonitis due to food and vomit, secondary malignant neoplasm of bone/bone marrow, malignant neoplasm of lung, pneumonia, congestive heart failure, and acute renal failure. We identified the parent ICD code for each of these, i.e., the first three characters of the ICD-10 code (such as C78 for secondary malignant neoplasm of lung) and checked whether there was any code starting with that present in the history of a patient. The variables and the prevalence of them in our cohort is shown in Table S2. We included these into our initial MAGGIC set, subsequently with some feature engineering. This included combining the use of beta-blockers, and angiotensin-converting-enzyme (ACE) inhibitors/angiotensin receptor blockers (ARBs) (named “num_antihypertensives”) and the cancers (identified by the explainability analysis on the transformer model – named “num_cancer”). This means if the patient does not have any cancer history, “num_cancer” would be 0, 1 if they have one of these cancers, and so on. Similarly for the anti-hypertensives as well.

Finally, to build a truly parsimonious model (as currently we were using 21 variables), we conducted SHAP analysis to identify the most predictive variables along with including year of birth as a variable.^3^ The reason for including year of birth was to use it as a proxy of birth cohort effects, that have shown to effect populations born in different years having different distribution of several diseases (such as colorectal cancer and atrial fibrillation) ^4-6^. The final model included 11 variables: year of birth, whether HF occurred 18 months before baseline, age, cancer history (three different classes of cancers), prescription history of anti-hypertensives (beta-blockers, ACE Inhibitors/ARBs), BMI, history of acute renal failure, pneumonia, cardiac arrest, respiratory failure, and chronic obstructive pulmonary disease (COPD).

For all models, the predictors were captured from both primary care and HES records. Co-morbidities and medication use was extracted using established and curated phenotyping dictionaries for CPRD Aurum. For vital signs, such as systolic blood pressure, creatinine, and body mass index, average of records that were collected in the 36 months leading up to baseline were used as baseline variables for modelling. Last known smoking status in the 36 months leading up to baseline was used as baseline smoking status.

#### AI Modelling:

MLP models utilise the Scalable Continuous-Time Survival Model through Ordinary Differential Equation Networks (SODEN) framework for modelling. Instead of maximum likelihood estimation (MLE) frameworks that utilise costly integral calculations for model training on censored data, the SODEN framework alternatively poses MLE as a differential-equation constrained optimisation objective.^7^ Furthermore, unlike previous proportional hazard frameworks that have not been shown to be theoretically robust for stochastic gradient descent (SGD) using mini-batching (i.e., random non-overlapping subsets of the derivation dataset), the SODEN framework alleviates many issues of established survival model frameworks that have trouble scaling to “big data”. Specifically, with ordinary differential equations modelling the time-to-event distribution, the framework for MLE on censored data becomes more flexible (i.e., absent of strong structural assumptions of the shape of the survival/hazard distribution), more appropriately scalable (i.e., can be trained using SGD).

Standard training protocols were implemented for all MLP models, including batch normalisation to stabilise learning, dropout for regularisation, and an early stopping criterion guided by the concordance index (C-index) on a held-out portion of the derivation dataset to prevent overfitting.

The different models underwent a development process involved iterative feature engineering and regularisation techniques to optimise performance and promote parsimony (**Figure S2**). The steps included:

1. Baseline Model (MLP-MAGGIC): We began by developing a baseline model using the established MAGGIC predictor set. This initial model, named MLP-MAGGIC, served as a benchmark for subsequent enhancements.
2. Enriched Model (MLP-Explainability): To improve predictive power, we augmented the feature set. While our primary goal was parsimony in the number of features for clinical interpretability and ease of use, we were informed by findings from more complex models, specifically the Transformer-based TRisk model. This research highlighted the prognostic value of previously under-appreciated variables, such as specific cancer histories and renal failure. We also incorporated the patient's year of birth as a proxy for birth cohort effects This created an enriched model with 21 variables, named MLP-Explainability. (**Table S2** for the exact explainability variables and their prevalence in the CPRD cohort)
3. Final Distilled Model (SIMPLE-HF): As a final step to enforce parsimony, we used SHapley Additive exPlanations (SHAP)^24^ to analyse the MLP-Explainability model and distil its 21 features down to only the most impactful predictors. This process resulted in our final model, SIMPLE-HF (Simplified Intelligent Mortality Prediction for Longitudinal EHRs in HF patients), which uses a refined set of just 11 variables: year of birth, occurrence of HF 18 months before baseline, age, cancer history, prescription of beta-blockers or ACE inhibitors/ARBs, body mass index (BMI), and history of acute renal failure (**Figure S3** for results of SHAP analysis).

#### SHAP Analyses:

To create a parsimonious model, SHAP analysis was conducted on the full 21 variable set (MAGGIC + Explainability + year of birth). The analysis was done for the prediction of all-cause mortality as that was our primary outcome. The top 11 variables were chosen as input to our SIMPLE-HF model for prediction of both primary and secondary outcomes. The reason behind selecting the top 11 features was due to diminishing returns after adding more features in terms of discriminatory performance of the model. Figure S3 shows the results of this SHAP analyses.

#### Implementation of MAGGIC-EHR Models

Missing values for systolic blood pressure, smoking status, body mass index, creatinine, and NYHA Functional Class were imputed for the MAGGIC-EHR model. Procedurally, for imputation of all-cause mortality prediction MAGGIC-EHR model, all variables for each model were included for imputation along with Nelson-Aalen estimate of baseline cumulative hazards for the outcome of mortality. Imputations were conducted on each derivation and validation datasets of our cohort separately using the “mice: Multivariate Imputation by Chained Equations” package in R.^8^ Following the extraction of derivation and validation datasets, the statistical models were fit on each of the five imputed derivation datasets with appropriate predictors (i.e., 5 MAGGIC-EHR models) for prediction of the primary outcome, all-cause mortality. The five Cox models were pooled using Rubin’s rules and evaluated on the 5 imputed validation datasets. The predictions were also appropriately pooled using extensions of the Rubin’s rules (i.e., complementary log-log transformations) and utilised for downstream analysis. The same procedures for model fitting and evaluation on validation datasets were repeated for the analyses of other outcomes: (1) fatal and non-fatal cardiovascular event prediction, and (2) rehospitalisation prediction

It must be noted that the original MAGGIC model derivation study has solely investigated 12- and 36-month all-cause mortality.^9^ To serve as the benchmark conventional model in this work, we used MAGGIC-EHR to not only predict 12- and 36-month all-cause mortality but also to demonstrate benchmark performance for the secondary outcomes investigated. Interestingly, while MAGGIC was not derived for secondary outcome investigations, validation studies have indeed been conducted for other outcomes such as CV-related mortality^10^

### Supplementary Figures

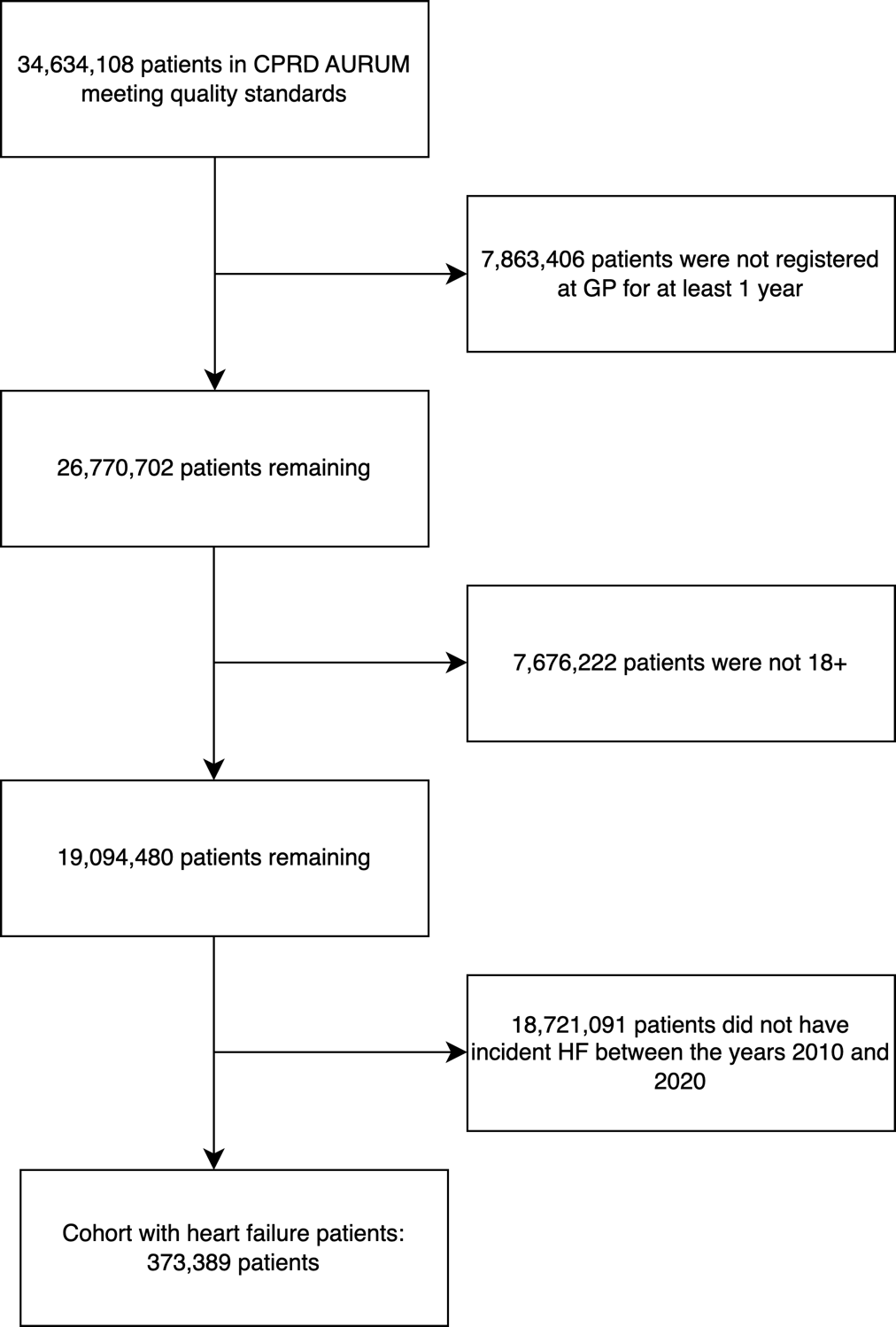

#### Figure S1. Cohort selection flowchart for Clinical Practice Research Datalink (CPRD) Aurum dataset.

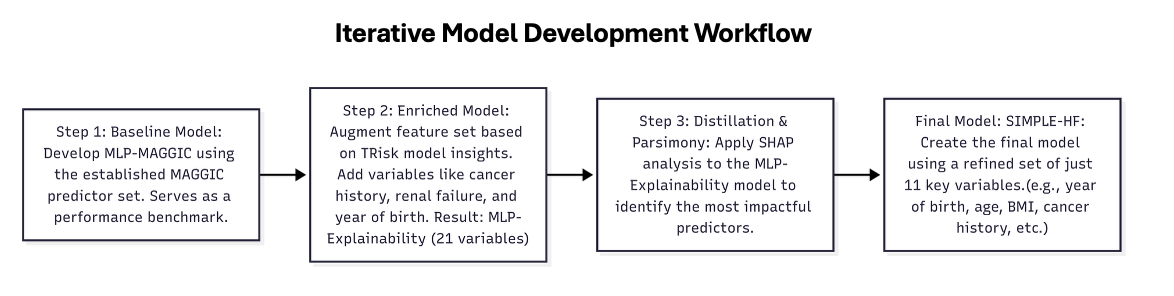

#### Figure S2: Iterative Model Development Workflow for the SIMPLE-HF Model.

*A schematic outlining the iterative model development process of SIMPLE-HF. The workflow begins with a baseline model (MLP-MAGGIC) and is enhanced into an enriched model with 21 variables (MLP-Explainability). SHAP analysis is then applied to distil the most impactful predictors, resulting in the final, parsimonious SIMPLE-HF model that uses a refined set of 11 key variables.*

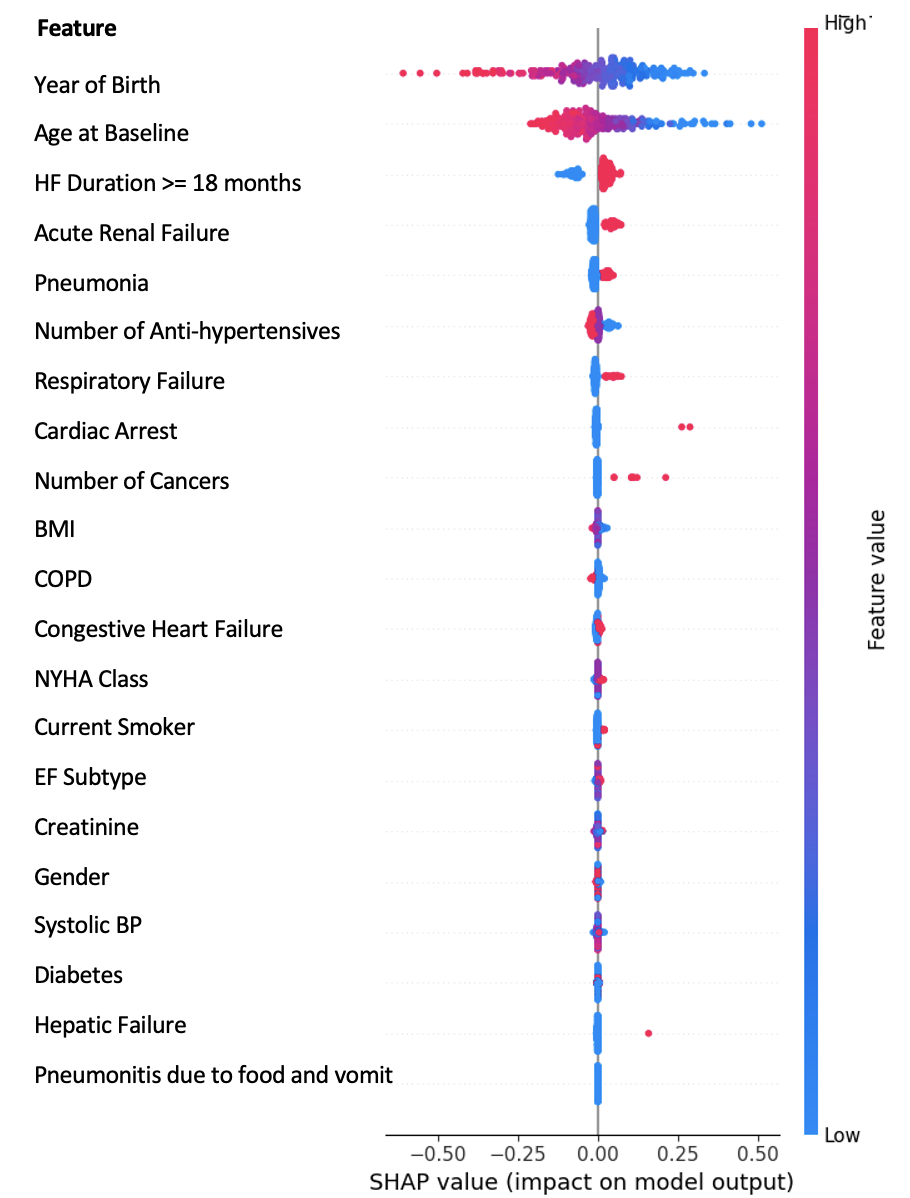

#### Figure S3: Global explanation of the MLP-Explainability model for mortality prediction using SHAP values

*SHAP summary plot displaying the marginal contribution of 21 baseline clinical variables to the model’s predicted probability of all-cause mortality. Features are ordered by their mean absolute SHAP value (overall predictive value).*

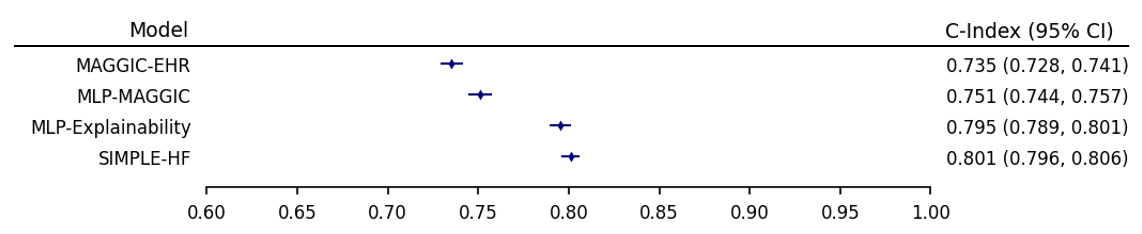

#### Figure S4: Discriminative performance for 12-month risk prediction of all-cause mortality in UK validation data

*Discrimination is provided in this forest plot as assessed by C-index with 95% confidence intervals (CI)*

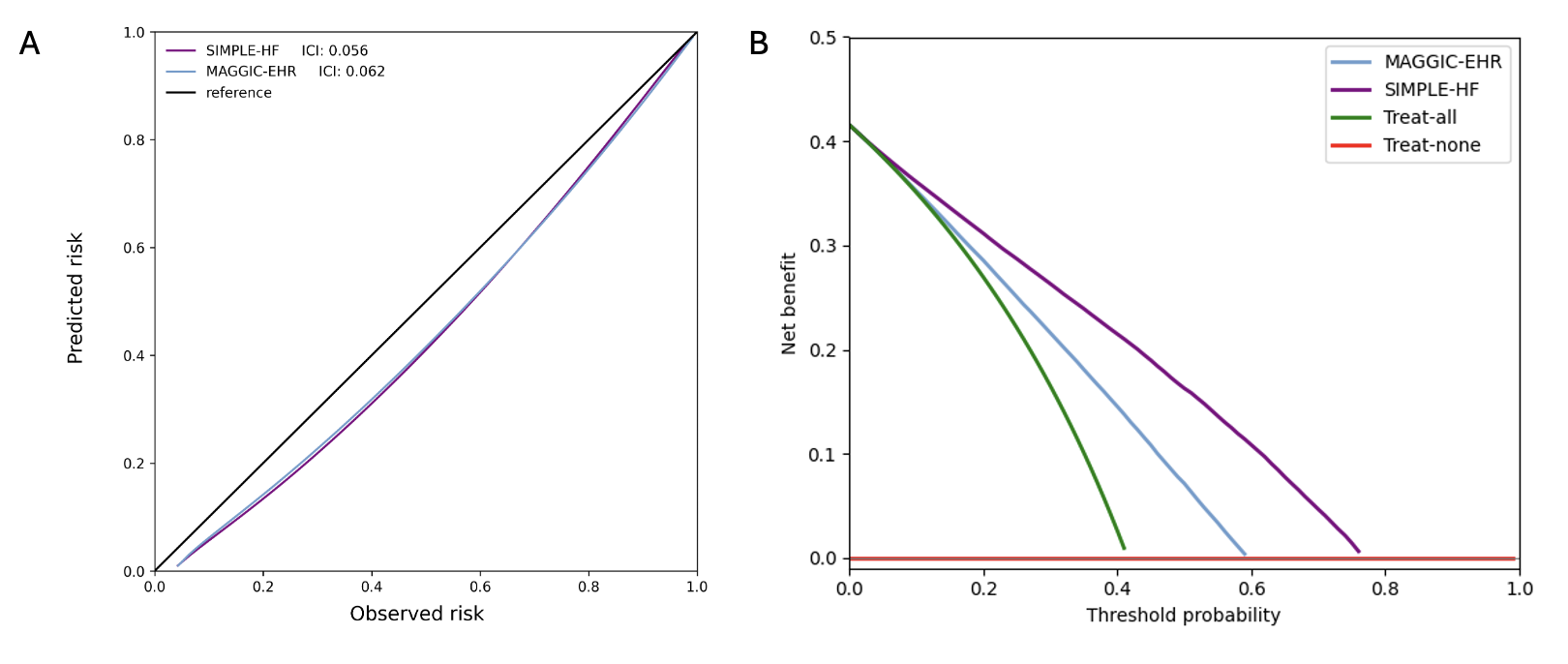

#### Figure S5: Calibration curves, and net benefit analysis for 36-month risk prediction of all-cause mortality in UK validation data

*Calibration curves with integrated calibration indices (ICI) (A), and decision curve analysis (B) are presented for all-cause mortality at 36-month timepoint. For (B) ICI, lower is better with reference (black line) presenting optimal ICI of 0.0.*

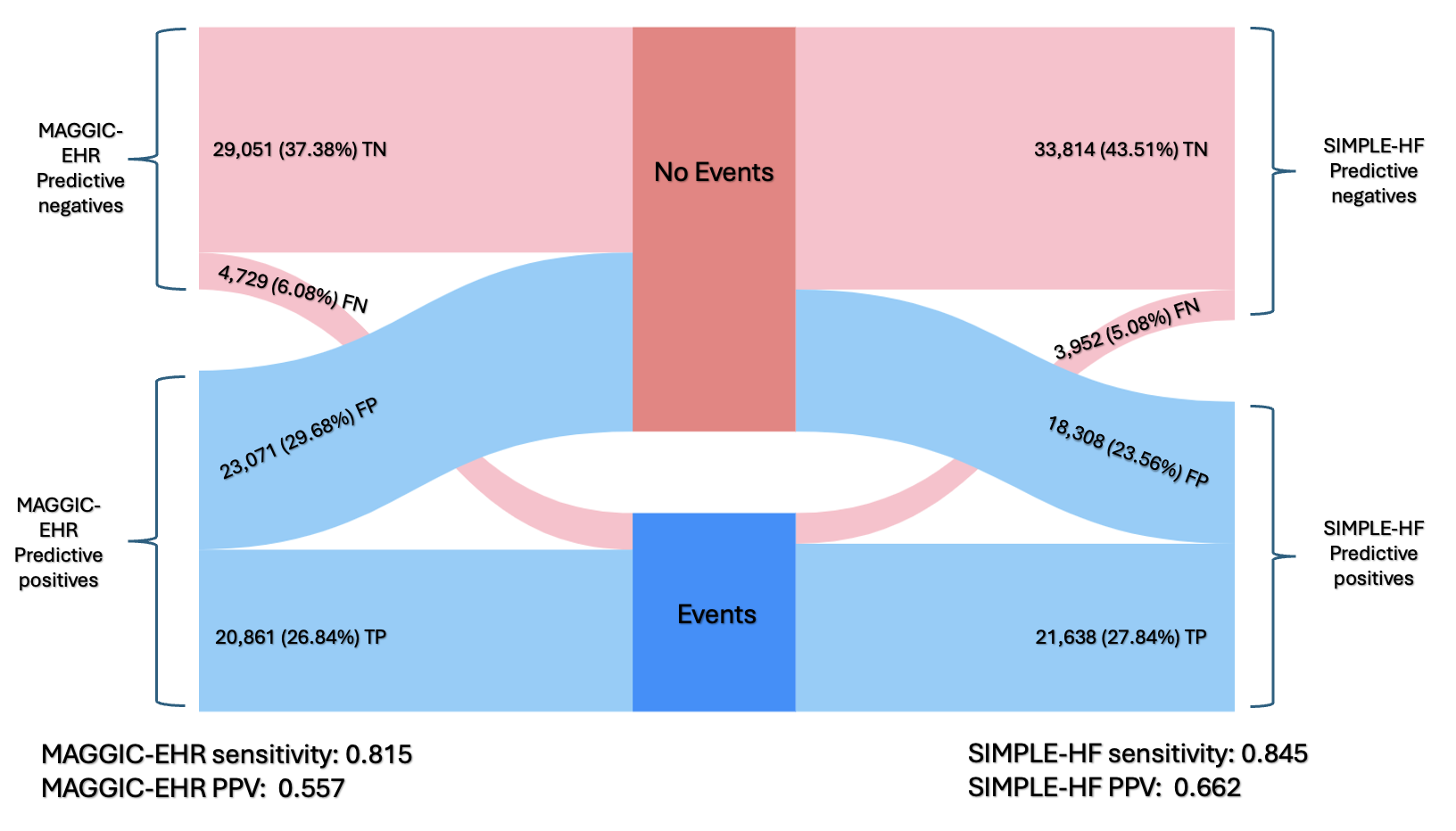

#### Figure S6: Impact analyses at the 50% decision threshold for 36- month all-cause mortality prediction on UK validation data.

*Sankey diagrams compare predicted outcomes between the models, showing how patient classification compares to actual outcomes, "Events" (i.e., death) and "No events " categories at 50% threshold (denoted as dark blue and red respectively). TP: true positive; TN: true negative; FP: false positive; FN: false negatives.*

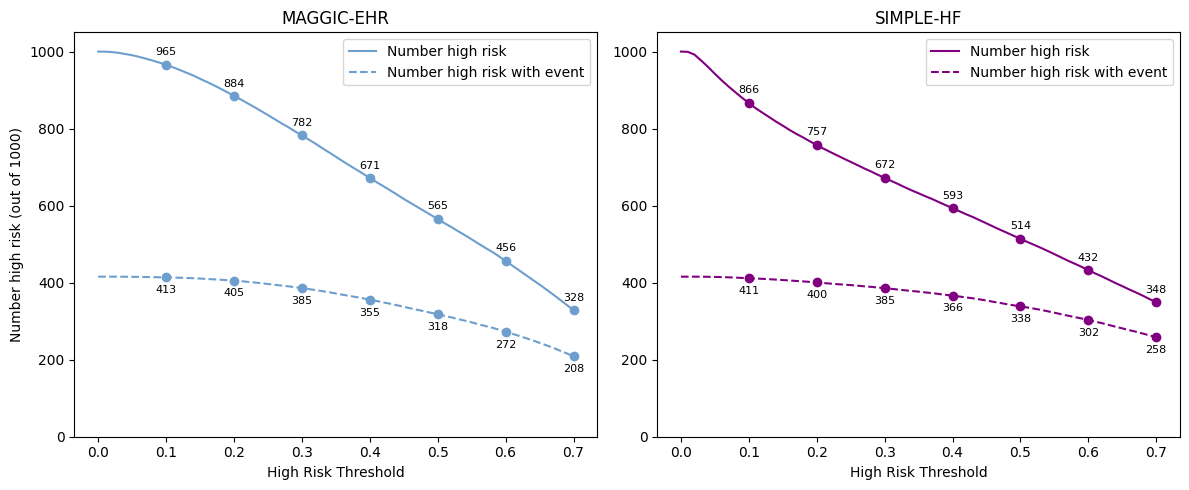

*Figure S7: Clinical-impact curves for 36-month all-cause mortality prediction in the UK validation cohort.*
*Comparison of MAGGIC-EHR (right) with the SIMPLE-HF (left). For each possible decision threshold on the x-axis, the solid line represents the number of patients per 1000 who would be classified as high risk, whereas the dashed line represents the subset of those high-risk patients who experienced the outcome (death). Numeric labels denote the corresponding absolute counts,*

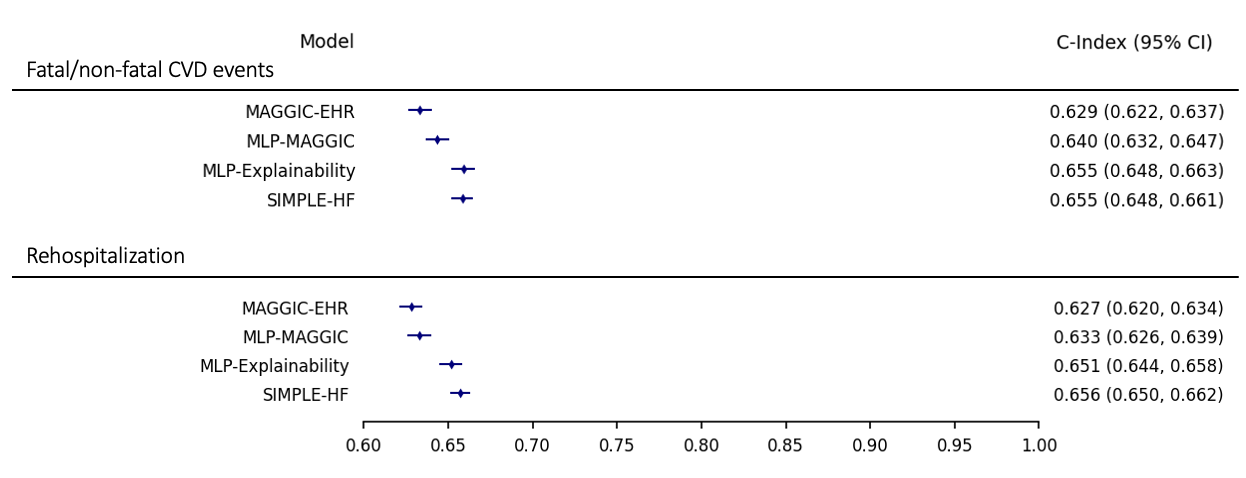

*Figure S8: Discriminative performance of models for 12-month risk prediction of various outcomes on UK validation data.*

*Discrimination is provided in this forest plot as assessed by C-index with 95% confidence intervals (CI) for various outcome investigations at 12-month timepoint; CVD: cardiovascular.*

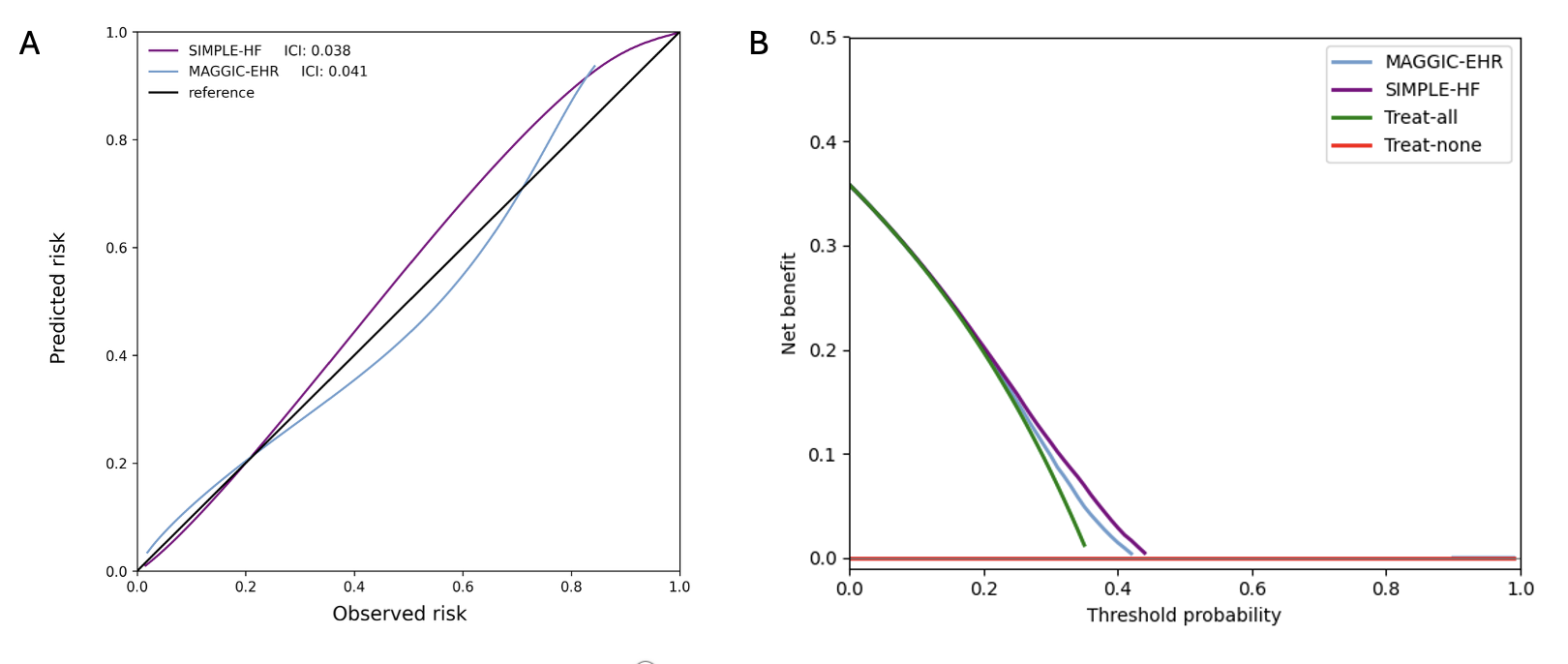

#### Figure S9: Calibration curves, and net benefit analysis for 12-month risk prediction of fatal/non-fatal cardiovascular events in UK validation data

*Calibration curves with integrated calibration indices (ICI) (A), and decision curve analysis (B) are presented for fatal/non-fatal cardiovascular events at 12-month timepoint. For (B) ICI, lower is better with reference (black line) presenting optimal ICI of 0.0.*

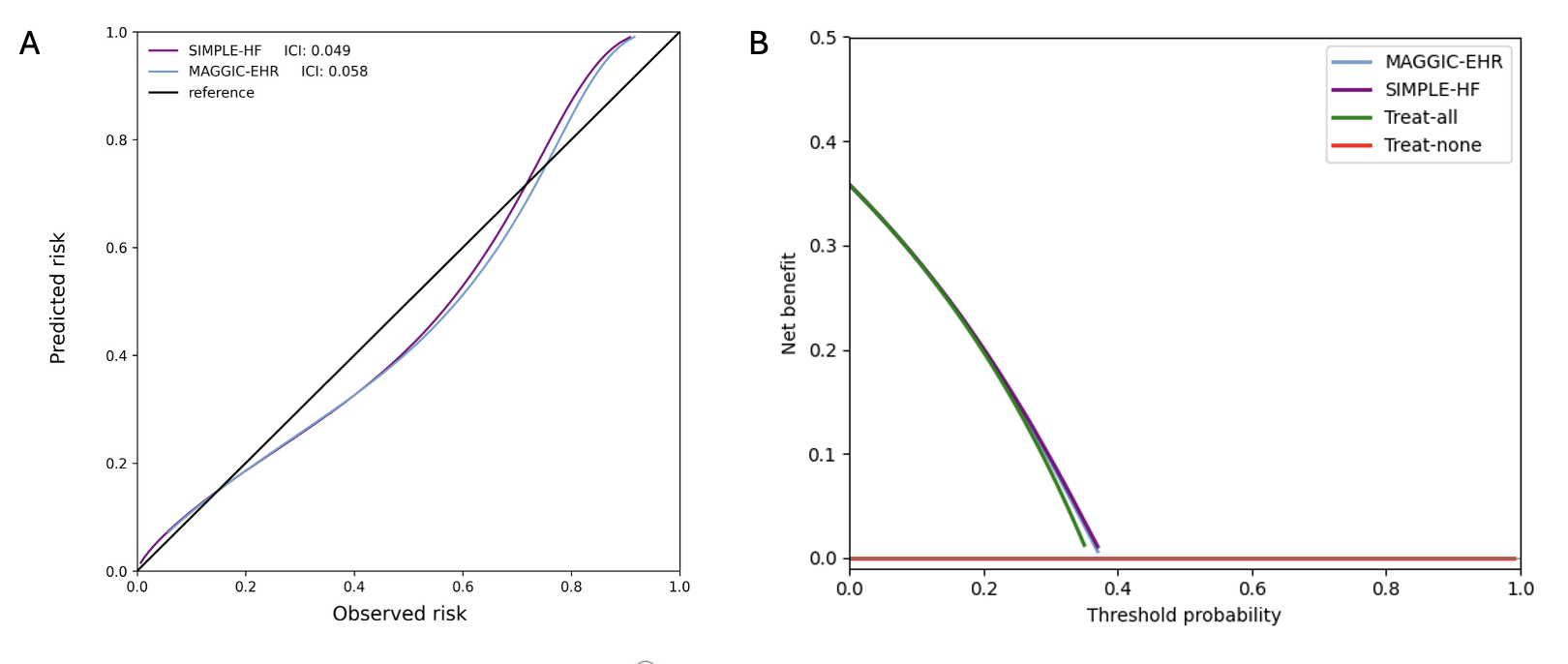

#### Figure S10: Calibration curves, and net benefit analysis for 36-month risk prediction of fatal/non-fatal cardiovascular events in UK validation data

*Calibration curves with integrated calibration indices (ICI) (A), and decision curve analysis (B) are presented for fatal/non-fatal cardiovascular events at 36-month timepoint. For (B) ICI, lower is better with reference (black line) presenting optimal ICI of 0.0.*

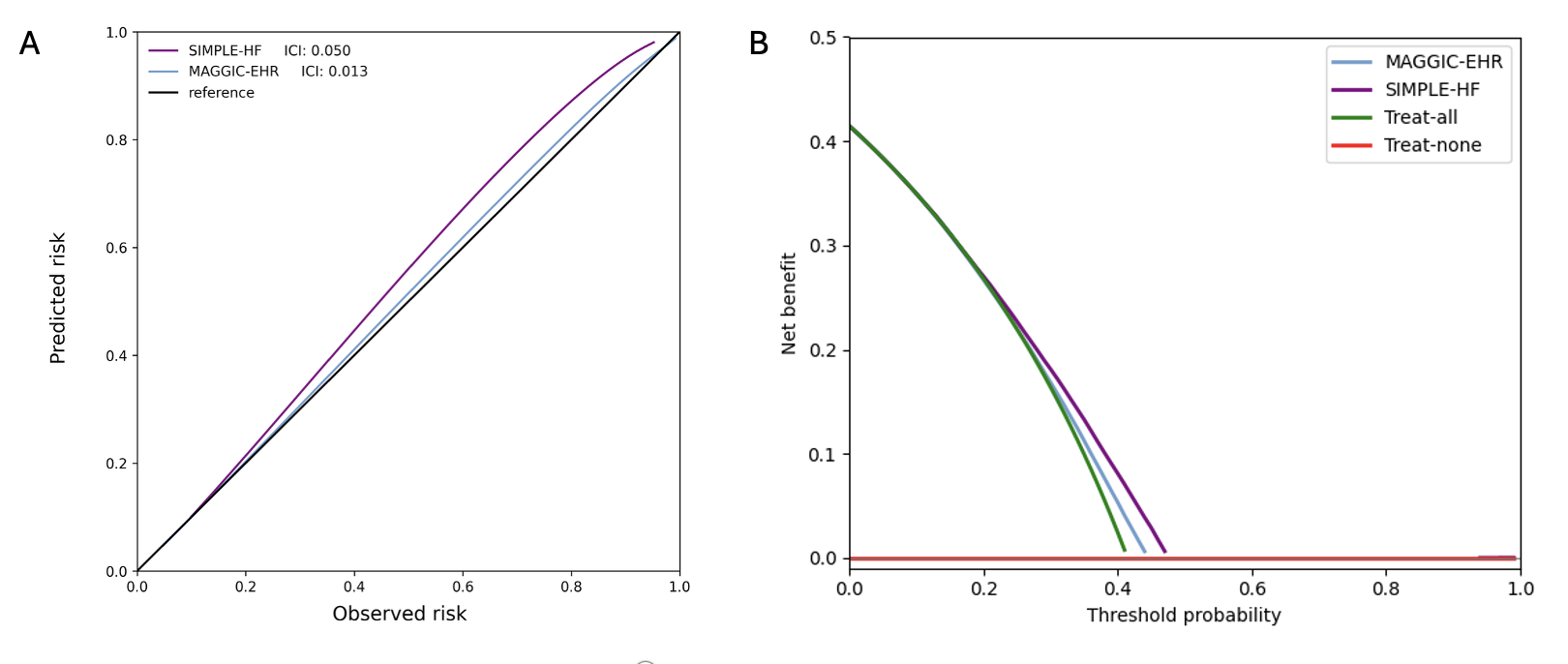

#### Figure S11: Calibration curves, and net benefit analysis for 12-month risk prediction of rehospitalization in UK validation data

*Calibration curves with integrated calibration indices (ICI) (A), and decision curve analysis (B) are presented for rehospitalization at 12-month timepoint. For (B) ICI, lower is better with reference (black line) presenting optimal ICI of 0.0.*

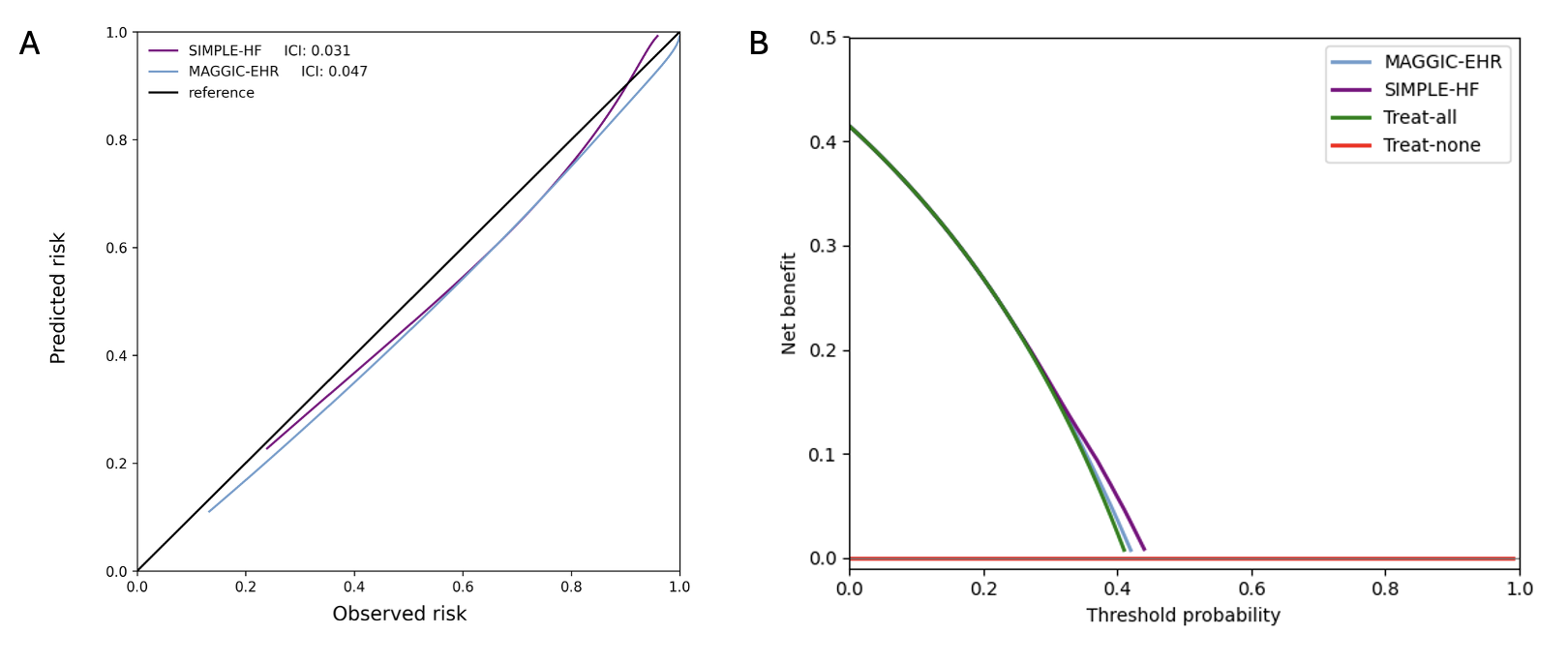

#### Figure S12: Calibration curves, and net benefit analysis for 36-month risk prediction of rehospitalization in UK validation data

*Calibration curves with integrated calibration indices (ICI) (A), and decision curve analysis (B) are presented for rehospitalization at 36-month timepoint. For (B) ICI, lower is better with reference (black line) presenting optimal ICI of 0.0.*

### Supplementary Tables

#### Table S1. Model hyperparameters and other settings for MLP models

| **Hyperparameters** | **Value** |
| --- | --- |
| Number of hidden layers | 3 |
| Hidden layer sizes | [64,128,64] |
| Hidden activation function | Leaky Rectified Linear Unit (RELU) function |
| Learning Rate (with scheduler) | 0.005 |
| Weight decay | 1e-5 |

#### Table S2. Features identified by TRisk and their prevalence in UK cohort

| **Medical Encounter** | **Prevalence (%)** |
| --- | --- |
| Cardiac Arrest | 0.82 |
| Secondary malignant Neoplasm Lung | 1.45 |
| Hepatic Failure | 0.79 |
| Respiratory Failure | 10.42 |
| Pneumonitis due to food and vomit | 0.06 |
| Secondary malignant neoplasm bone marrow | 1.89 |
| Malignant Neoplasm Lung | 1.57 |
| Pneumonia | 29.45 |
| Congestive Heart Failure | 43.65 |
| Acute Renal Failure | 26.13 |

#### Table S3. Area under the precision-recall curve (AUPRC) metrics for all models for all 12-month risk prediction investigations on UK validation data

|  | | AUPRC (95% CI) |
| --- | --- | --- |
| All-cause mortality | MAGGIC-EHR | 0.548 (0.541, 555) |
|  | MLP-MAGGIC | 0.560 (0.553, 0.567) |
|  | MLP-Explainability | 0.643 (0.636, 0.650) |
|  | SIMPLE-HF | 0.684 (0.677, 0.691) |
| Fatal and non-fatal cardiovascular events | MAGGIC-EHR | 0.371 (0.363; 0.378) |
|  | MLP-MAGGIC | 0.378 (0.371; 0.386) |
|  | MLP-Explainability | 0.376 (0.368; 0.383) |
|  | SIMPLE-HF | 0.390 (0.383; 0.398) |
| Rehospitalization | MAGGIC-EHR | 0.368 (0.361; 0.375) |
|  | MLP-MAGGIC | 0.369 (0.362; 0.376) |
|  | MLP-Explainability | 0.367 (0.360; 0.374) |
|  | SIMPLE-HF | 0.368 (0.361; 0.375) |

#### Table S4. Integrated calibration index (ICI) for various 12-month risk prediction investigations across all models on UK validation data

| Integrated Calibration Index (ICI)* | | | |
| --- | --- | --- | --- |
|  | All-cause mortality | Non-fatal/fatal cardiovascular events | Rehospitalization |
| MAGGIC-EHR | 0.038 | 0.041 | 0.013 |
| MLP-MAGGIC | 0.025 | 0.036 | 0.012 |
| MLP-Explainability | 0.034 | 0.030 | 0.037 |
| SIMPLE-HF | 0.037 | 0.038 | 0.050 |

**lower is better*

#### Table S5. Integrated calibration index (ICI) for various 36-month risk prediction investigations across all models on UK validation data

| Integrated Calibration Index (ICI)* | | | |
| --- | --- | --- | --- |
|  | All-cause mortality | Non-fatal/fatal cardiovascular events | Rehospitalization |
| MAGGIC-EHR | 0.062 | 0.058 | 0.047 |
| MLP-MAGGIC | 0.100 | 0.048 | 0.089 |
| MLP-Explainability | 0.087 | 0.124 | 0.078 |
| SIMPLE-HF | 0.056 | 0.049 | 0.031 |

**lower is better*

#### Table S6. Impact analyses at the 50% decision threshold for 12- and 36-month all-cause mortality prediction on UK validation data

| Timepoint | Models |  |  | Metrics | | | | |
| --- | --- | --- | --- | --- | --- | --- | --- | --- |
|  |  |  | PP | TP | FP | FN | PPV | Sensitivity |
| 12-month | MAGGIC-EHR |  | 21,303 | 12,147 | 9,156 | 13,443 | 0.570 | 0.475 |
|  | SIMPLE-HF |  | 26,412 | 16,965 | 9,447 | 8,625 | 0.642 | 0.663 |
| 36-month | MAGGIC-EHR |  | 43,932 | 20,861 | 23,071 | 4,729 | 0.557 | 0.815 |
|  | SIMPLE-HF |  | 39,946 | 21,638 | 18,308 | 3.952 | 0.662 | 0.845 |

*PP: predictive positive; TP: true positives; FP: false positives; FN: false negatives; PPV: positive predictive value*
